# The Cleaning Simulation: Applying Predictive Decision Trees for Chemical Exposure Risks and Asthma-Like Symptoms in Laboratory Workers

**DOI:** 10.1101/2025.11.25.25341001

**Authors:** Hayden D. Hedman

## Abstract

Exposure to chemical irritants in laboratory and medical environments poses significant health risks to workers, particularly in relation to asthma-like symptoms. Routine cleaning practices, which often involve the use of strong chemical agents to maintain hygienic settings, have been shown to contribute to respiratory issues. Laboratories, where chemicals such as hydrochloric acid and ammonia are frequently used, represent an underexplored context in the study of occupational asthma. While much of the research on chemical exposure has focused on industrial and high-risk occupations or large cohort populations, less attention has been given to the less obvious risks in laboratory and medical environments. Given the growing reliance on cleaning agents to maintain sterile and safe workspaces in scientific research and healthcare facilities, I find this gap particularly concerning. In my study, I used a simulated cohort based on key demographic and exposure patterns from foundational research to assess the impact of chemical exposure from cleaning products in laboratory environments. I applied four supervised machine learning models to evaluate the relationship between chemical exposures and asthma-like symptoms: (1) Decision Trees, (2) Random Forest, (3) Gradient Boosting, and (4) XGBoost. I found that high exposures to hydrochloric acid and ammonia were significantly associated with asthma-like symptoms, and workplace type also played a critical role in determining asthma risk. This research provides a data-driven framework for assessing and predicting asthma-like symptoms in workers exposed to cleaning agents and highlights the potential for integrating predictive modeling into occupational health and safety monitoring. Future work should explore dose-response relationships and the temporal dynamics of chemical exposure to further refine these models and better understand long-term health risks.

## 1. Introduction

Exposure to chemical irritants in occupational settings, especially in laboratory and medical environments *(1,2)*, poses substantial health risks to workers. Routine cleaning practices, which often involve the use of strong chemical agents to maintain hygienic settings *(3)*, have been shown to contribute to respiratory issues such as asthma-like symptoms *(4–7)*. Laboratories, where chemicals (e.g., hydrochloric acid and ammonia) are widely documented airway irritants *(8)*, yet most research on occupational asthma focuses on industrial settings or large, high-risk cohorts *(9,10)*. For laboratory personnel who engage in frequent cleaning activities or work in close proximity to these agents, this gap is concerning. Conditions such as work-related asthma are multifactorial and rare *(1,11)*, making subtle risks harder to detect without systematic analysis.

To address this gap, the present study examines how routine exposure to commonly used cleaning agents may contribute to asthma-like symptoms among laboratory and medical staff. Because real-world data on these exposures are limited for laboratory settings, I developed a simulated workforce cohort based on established occupational-exposure findings *(4)*, reflecting realistic demographic and exposure patterns. This approach enables a structured evaluation of respiratory risk in a setting where direct surveillance data are scarce.

Finally, this work uses a predictive modeling framework to explore how chemical exposures relate to respiratory symptoms and how such approaches might support future occupational health monitoring in laboratory environments. The primary aims are to: (1) estimate the relationship between chemical exposures from routine cleaning agents and asthma-like symptoms, (2) compare predictive performance across several modeling approaches, and (3) provide a framework for integrating data-driven tools into laboratory health and safety practices.

## 2. Materials and Methods

### 2.1. Adaptation of Vizcaya et al. (2011)

I developed the simulated cohort based on the distributions reported by Vizcaya et al. (2011) *(4)*, replicating key demographic, exposure, and behavioral characteristics (Table 1). Given the lack of real-world data on chemical exposure in laboratory environments, simulation was used to create a realistic model of the population. Sex was modeled to match the predominance of female cleaners (82% female), and age was simulated using the study’s central tendency (mean = 45 years; SD = 10). Smoking status followed the reported frequencies (30% current smokers; 10% former smokers, 60% never smokers). Although Vizcaya et al. did not specify the proportion of foreign-born workers, I included a 30% foreign-born proportion to reflect demographic patterns commonly reported in cleaning-industry workforces *(12,13)*. This variable was retained for completeness but contributed minimally to the modeled risk structure.

**Table 1.**
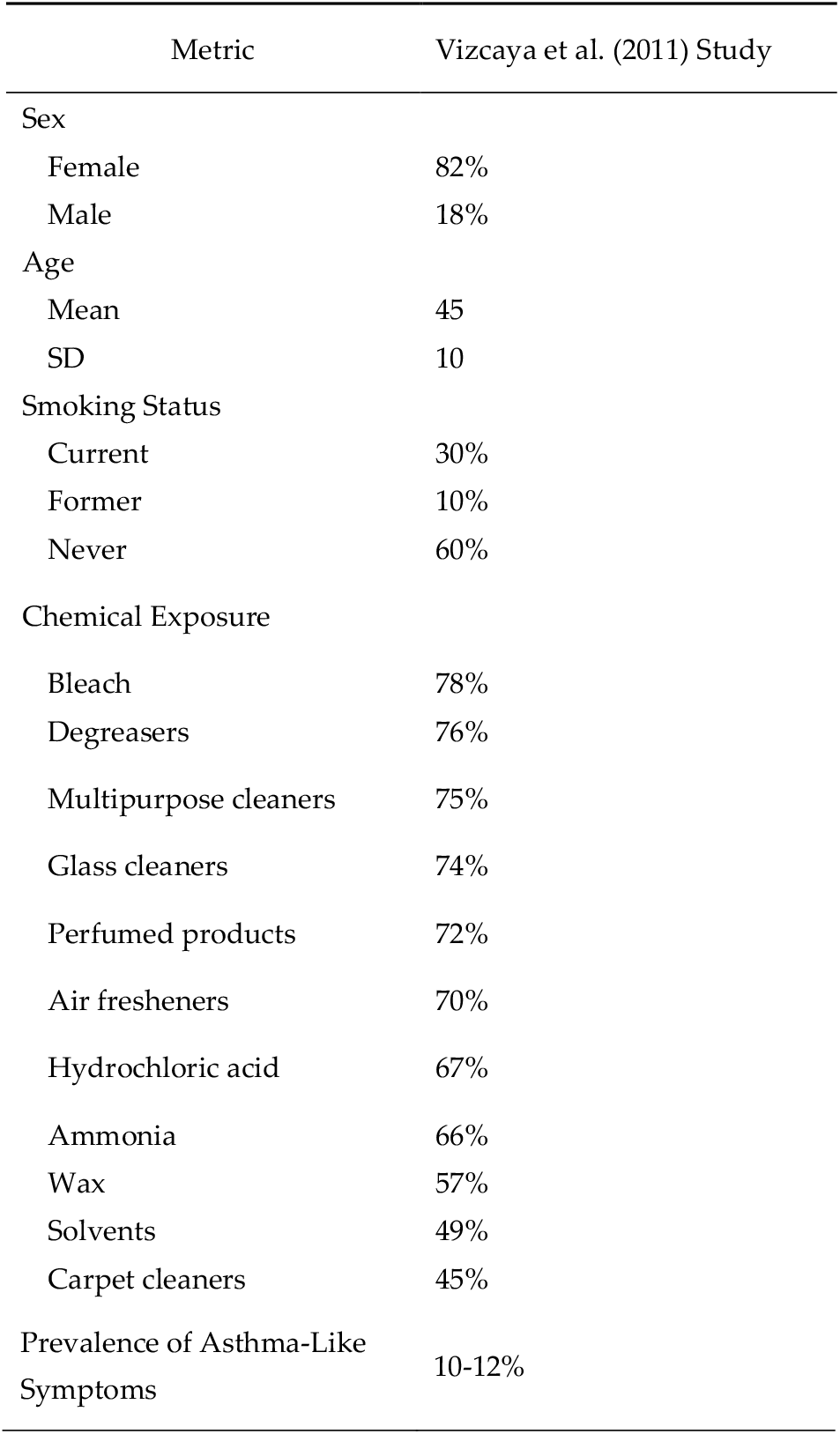
Comparison of key demographic, occupational, and exposure variables reported by Vizcaya et al. (2011) *(4)* and the corresponding values applied in the simulated cohort.

Workplace settings were simulated using the study’s distribution, ensuring proportional representation of hospital cleaners (24%) and common-area workers (25%). Chemical exposures including hydrochloric acid (10%), ammonia (15%), and degreasers (18%) were incorporated directly from the study’s irritant profile *(4)* (Table 1). To model asthma-like symptoms, I implemented a logistic risk function that integrated these exposure variables along with smoking and foreign-born status, calibrating the model to produce an overall symptom prevalence of approximately 11%, consistent with the original study. Although not every variable from Vizcaya and colleagues *(4)* could be directly mapped due to data constraints or relevance, the simulation captured the essential demographic, occupational, and chemical-exposure patterns needed to reflect realistic environmental conditions and worker characteristics most relevant to evaluating respiratory risk within cleaning and laboratory support environments.

### 2.2. Simulated Data Generation and Exposure Modeling

I generated a synthetic dataset in Python (version 3.11.9) using NumPy and pandas to replicate the demographic, workplace, and chemical-exposure patterns from Vizcaya et al. (2011) (Table 1). All variable distributions, including sex, smoking status, workplace type, and chemical exposures, were parameterized based on the proportions reported in the study, with no additional assumptions beyond those outlined in Section 2.1. I simulated 20,000 observations to ensure sufficient sample size for downstream modeling. Categorical variables were generated using multinomial or binomial sampling, while continuous variables (e.g., age) were simulated using truncated normal distributions.

Asthma-like symptoms were modeled using a logistic regression function calibrated to the symptom prevalence target (~11%) from the source study. Feature associations were assessed using Chi-square tests for categorical variables and Cohen’s d for continuous variables, as described in Section 3.1.

### 2.3. Classification Modeling for Predicting Asthma-Like Symptoms

In the third stage of the analysis, I developed a supervised classification pipeline in Python to predict asthma-like symptoms using the simulated cohort adapted from Vizcaya et al. (2011) *(4)*. I evaluated four tree-based model types commonly used in applied classification research *(14,15)*: (1) Decision Trees (DT), which classify individuals through a sequence of simple, rule-based splits and offer high interpretability *(16)* (cite); (2) Random Forests (RF), which aggregate predictions across multiple decision trees to improve stability and reduce overfitting *(17)*; (3) Gradient Boosting (GB), which builds trees sequentially so that each tree corrects errors made by prior trees *(18)*; and (4) XGBoost (Extreme Gradient Boosting) *(19)*, an efficient gradient-boosting implementation widely used when predictor–outcome relationships are nonlinear or involve interacting risk factors *(20,21)*.

To better represent occupational and chemical risk patterns, I constructed several domain-informed features prior to model training (Table 2). These included (1) a cumulative chemical-exposure score (total number of high-exposure products), (2) a binary indicator for workers exposed to multiple high-frequency irritants, (3) an interaction term between smoking status and workplace type to capture combined behavioral and occupational risk, and (4) an indicator for potentially vulnerable workers (foreign-born individuals with elevated chemical exposure). Categorical variables (sex, smoking category, work-place) were one-hot encoded, continuous variables (age and cumulative exposure) were standardized, and binary chemical-exposure indicators were retained without transformation.

**Table 2.** Demographic, workplace, and exposure characteristics of the simulated dataset, including the observed frequency of asthma-like symptoms.

| Metric | Observed Value |
| --- | --- |
| Sex |  |
| Female | 82.25% |
| Male | 17.75% |
| Age (years) |  |
| Mean | 44.93 |
| SD | 9.95 |
| Smoking status |  |
| Never | 60.06% |
| Former | 9.83% |
| Current | 30.12% |
| Foreign-born | 30.16% |
| Workplace type |  |
| Home | 28.18% |
| Common areas | 24.93% |
| Hospital | 23.78% |
| School | 4.96% |
| Other healthcare | 3.21% |
| Chemical Exposures |  |
| Hydrochloric acid | 10.18% |
| Ammonia | 15.46% |
| Degreasers | 18.09% |
| Multipurpose cleaners | 39.44% |
| Wax | 10.36% |
| Asthma-like symptoms | 5.12% |

Because asthma-like symptoms were relatively uncommon in the simulated cohort (5.1%), I observed substantial class imbalance during exploratory analysis. To address this, I applied SMOTE (Synthetic Minority Oversampling Technique) *(22)* to the training sets for the DT, RF, and GB models. SMOTE generated additional synthetic examples of symptomatic workers, improving the models’ ability to learn patterns associated with respiratory irritation. For XGBoost, oversampling was not required; instead, I used its internal class-imbalance weighting parameter (scale_pos_weight) *(23)*, which upweighted symptomatic cases during training without altering the data structure.

For model development, I partitioned the dataset into 70% training, 15% validation, and 15% testing, using stratified sampling to maintain the underlying outcome proportion. This approach minimized overfitting and provided an unbiased assessment of each model’s ability to identify symptomatic workers using data not observed during training. Model performance was evaluated using several standard metrics: accuracy, precision, recall (sensitivity), F1 score, AUC-ROC, and AUC-PR, with emphasis on AUC-PR given the imbalanced outcome distribution. Because false negatives were the primary concern from a laboratory and workplace-safety perspective *(1,24)*, I used threshold tuning *(25)* on the validation set to select a decision threshold that achieved ≥70% recall, prioritizing the detection of symptomatic workers. Probability-based threshold selection follows the broader approach used in occupational exposure modeling studies *(26)*, where probabilistic predictions support risk-based decision-making

### 2.4. Reproducibility and Documentation

The full documentation and code used to generate the simulated dataset (SM1) and to implement the classification models (SM2) are provided in the Supplementary Materials. The simulation incorporated all relevant variables from Vizcaya et al. (2011) *(25)*, reflecting the key demographic, exposure, and risk factors critical to investigating asthma-like symptoms in occupational cleaning workers.

### 2.5. Generative AI Statement

For this project, I used Generative AI tools *(27)* for coding support, troubleshooting, and initial brainstorming. All outputs were carefully reviewed.

## 3. Results

### 3.1. Overview and Descriptive Statistics

Overall, the simulated dataset contained 20,000 observations and 10 predictor variables, with no missing data. Given the highly imbalanced nature of the dataset (5.12% positive cases), model performance was evaluated using multiple metrics, including ROC AUC and PR AUC, to assess how well each model handled the imbalance (1,024 observations; imbalance ratio ≈ 18.5:1). This proportion is consistent with previous studies demonstrating that asthma-like respiratory symptoms occur in a minority of adults in occupational settings *(5)* and within general population prevalence ranges reported in large epidemiologic surveys *(28)*. The class distribution remained stable across the train, validation, and test sets (5.12%, 5.13%, and 5.10%, respectively). The feature set included one continuous variable (age), six binary variables (e.g., smoking status, chemical exposure), and three categorical variables (e.g., sex, workplace exposure).

Additionally, I examined feature associations, observing significant differences between the classes for several binary predictors. High chemical exposures (e.g., HCl, ammonia, degreasers) were significantly more common among symptomatic cases (Chisquare test, p < 0.05). For continuous predictors, age showed a negligible difference between groups (Cohen’s d = 0.015), indicating minimal variation in age between symptomatic and non-symptomatic workers.

### 3.2. Model Performance across Decision Trees

#### 3.1.1. Overall Model Comparison

I compared the performance of four classifiers: Decision Tree (DT), Random Forest (RF), Gradient Boosting (GB), and XGBoost (XGB), using sensitivity, specificity, ROC AUC, and PR AUC as the evaluation metrics (Table 3; Figure 1). RF achieved the highest sensitivity (75.2%) with moderate specificity (49.3%). XGB showed the highest specificity (94.2%) but slightly lower sensitivity (71.3%). For overall discrimination, RF had the top ROC AUC (0.673), followed closely by XGB (0.672). DT had the weakest performance across all metrics (ROC AUC: 0.631; PR AUC: 0.096), indicating limited ability to separate symptomatic and non-symptomatic workers compared with the ensemble models.

**Table 3.**
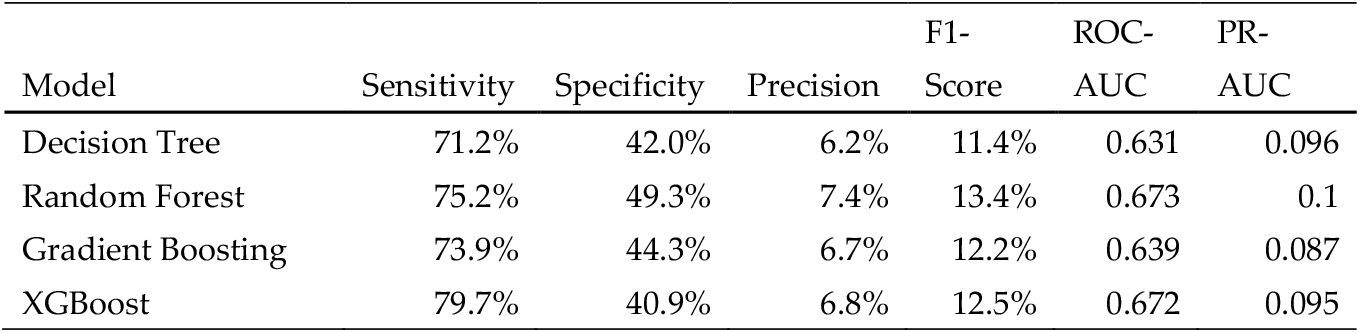
Performance comparison of four classifiers: Decision Tree (DT), Random Forest (RF), Gradient Boosting (GB), and XGBoost (XGB), based on sensitivity, specificity, precision, F1-score, ROC AUC, and PR AUC.

| Model | Sensitivity | Specificity | Precision | F1-Score | ROC-AUC | PR-AUC |
| --- | --- | --- | --- | --- | --- | --- |
| Decision Tree | 71.2% | 42.0% | 6.2% | 11.4% | 0.631 | 0.096 |
| Random Forest | 75.2% | 49.3% | 7.4% | 13.4% | 0.673 | 0.1 |
| Gradient Boosting | 73.9% | 44.3% | 6.7% | 12.2% | 0.639 | 0.087 |
| XGBoost | 79.7% | 40.9% | 6.8% | 12.5% | 0.672 | 0.095 |

**Figure 1.**
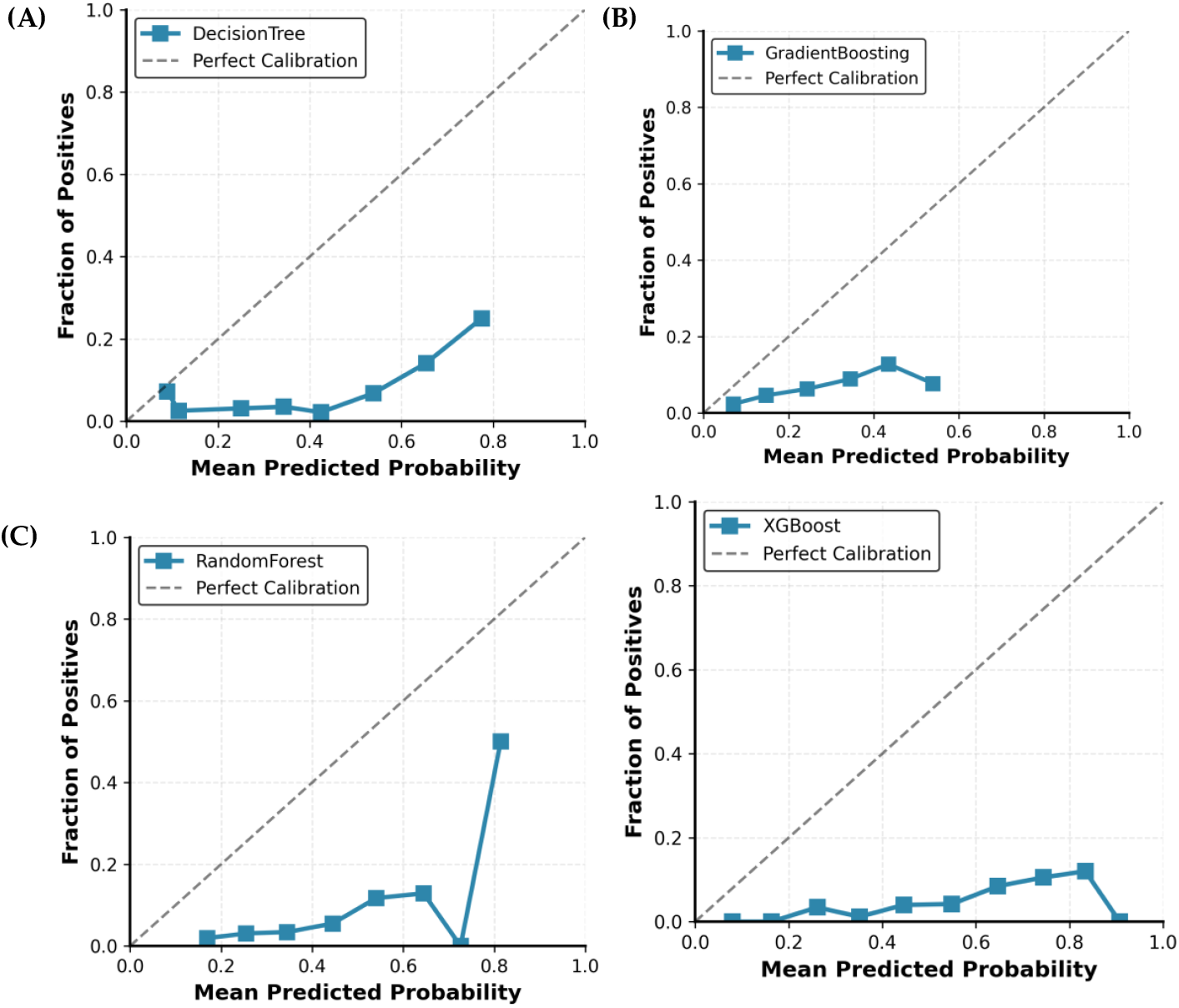
Calibration curves for each model: (A) Decision Tree, (B) Gradient Boosting, (C) Random Forest, and (D) XGBoost.

#### 3.1.2. Confusion Matrices

The confusion matrices further clarified the error patterns of each model, particularly the balance between true positives and false positives (Figure 1). RF correctly identified 115 positive cases out of 154 (sensitivity: 75.2%). DT identified 109 true positives out of 153 (sensitivity: 71.2%). XGB achieved the highest sensitivity (79.7%) but with lower specificity compared with RF (40.9% vs. 49.3%). These patterns highlight the tradeoff between sensitivity and specificity across models, with DT and GB producing higher false positive counts relative to RF and XGB.

#### 3.1.3. ROC Curves

Figure 2 compares the ROC curves for all models. RF achieved the highest ROC AUC (0.673), with XGB showing a nearly identical value (0.672). Although the AUC values were similar, RF maintained higher sensitivity across most thresholds. GB displayed moderate discrimination (ROC AUC: 0.639), and DT had the weakest performance (ROC AUC: 0.631). Overall, the ROC curves suggest that RF and XGB provided the strongest separation between positive and negative cases.

**Figure 2.**
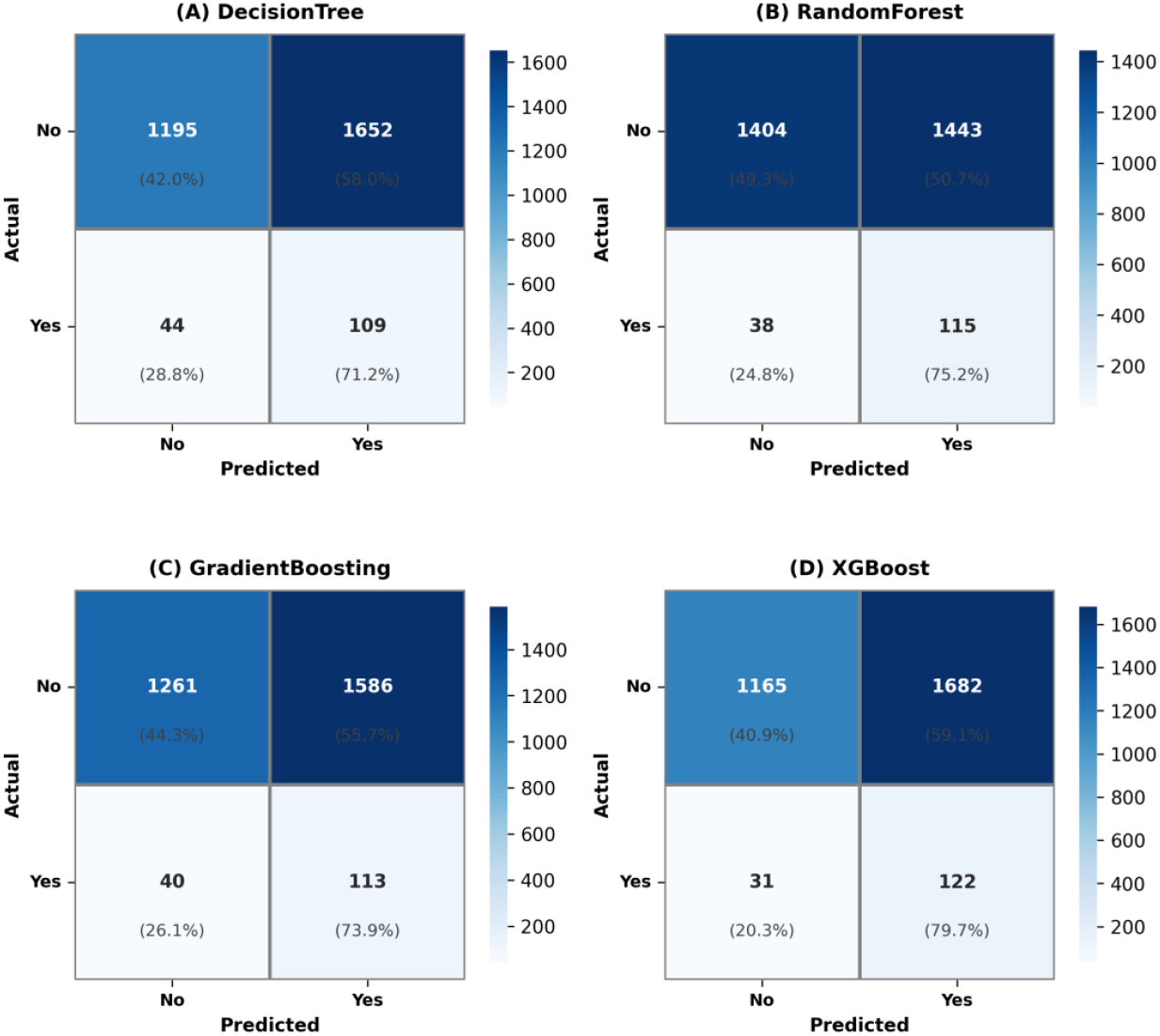
Confusion matrices for each model: (A) Decision Tree, (B) Random Forest, (C) Gradient Boosting, and (D) XGBoost.

#### 3.1.4. PR Curves

The precision–recall curves summarize how well each model identified positive cases under the high class imbalance (Figure 3). RF had the highest PR AUC (0.100), followed closely by XGB (0.095). Both scores indicate limited precision in this setting. GB and DT showed lower PR AUC values, consistent with their weaker performance in other metrics. Overall, the curves illustrate the difficulty of detecting positive cases and the low precision observed across all models.

**Figure 3.**
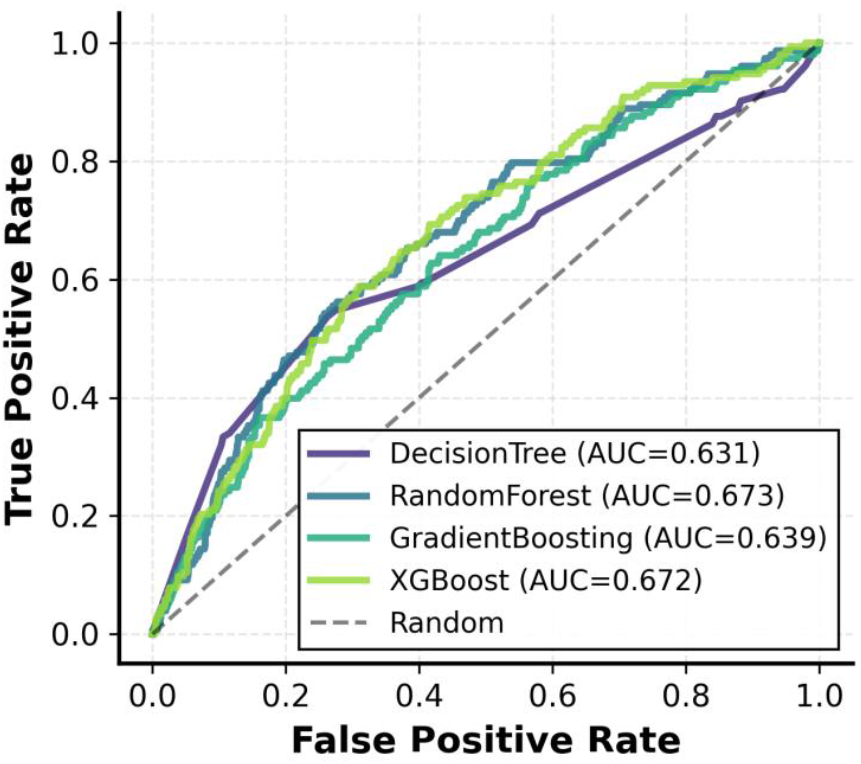
ROC curves for Decision Tree, Random Forest, Gradient Boosting, and XGBoost, with AUC values displayed for each model. The dashed line represents random performance (AUC = 0.5).

### 3.3. Model Reliability and Cross-Validation

In my cross-validation analysis, I observed clear differences in stability across the four models (Table 3; Figure 4). RF showed consistent performance, with a mean ROC AUC of 0.603 and a mean PR AUC of 0.090. XGB demonstrated similar stability, with a mean ROC AUC of 0.610 and a mean PR AUC of 0.093. DT, in contrast, had lower and more variable performance (mean ROC AUC: 0.567; mean PR AUC: 0.071). GB was the least stable model, with a mean ROC AUC of 0.574. Overall, I found that RF and XGB provided the most consistent results across folds.

**Figure 4.**
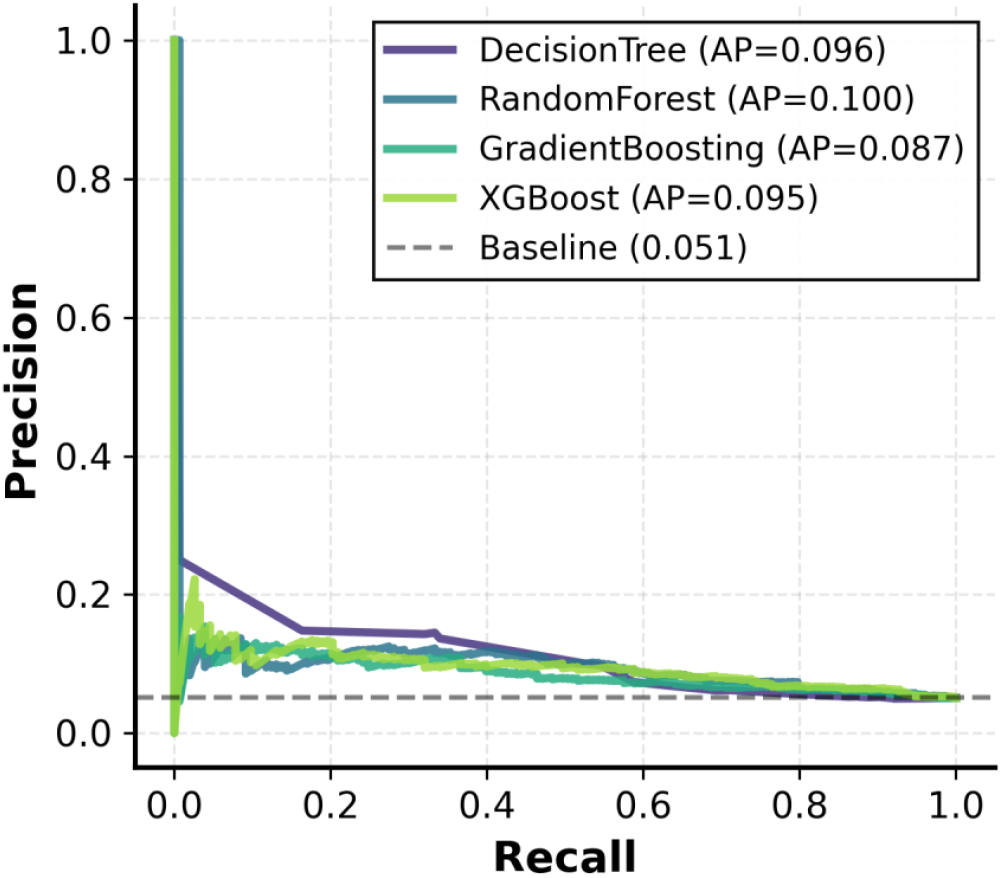
Precision-Recall curves for Decision Tree, Random Forest, Gradient Boosting, and XGBoost, with Average Precision (AP) values displayed for each model. The dashed line represents the baseline performance.

**Figure 5.**
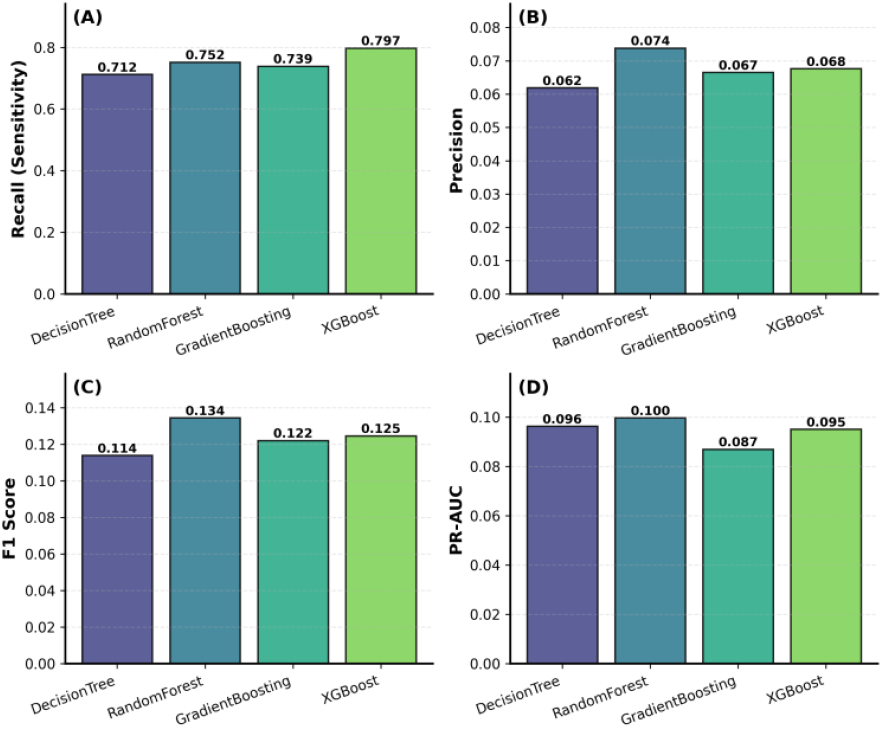
Comparison of model performance across multiple metrics: (A) Recall (Sensitivity), (B) Precision, (C) F1-Score, and (D) PR-AUC. The values for each model (Decision Tree, Random Forest, Gradient Boosting, and XGBoost) are displayed above the respective bars.

When I examined calibration, I noted substantial differences in the accuracy of predicted probabilities across models (Table 4). GB showed the best calibration with the lowest error (0.226). DT followed with a calibration error of 0.318. RF demonstrated moderate calibration accuracy (0.382), while XGB had the highest error (0.454), indicating poorer alignment between predicted and observed probabilities despite its stronger discrimination performance. These results led me to conclude that GB produced the most reliable probability estimates.

I also assessed the performance tradeoffs among the four models. XGB reached the highest ROC AUC (0.672) and PR AUC (0.095), but this came with lower specificity compared with RF. RF showed a more balanced profile overall, with higher sensitivity (75.2%), moderate specificity (49.3%), and the highest ROC AUC value (0.673). GB and DT produced lower sensitivity, precision, and PR AUC values, consistent with their weaker performance in earlier analyses. Taken together, I found that RF offered the best balance of sensitivity, specificity, and discrimination in identifying asthma-like symptoms.

## 4. Discussion

This study simulated the impact of common cleaning products in laboratory and medical environments on the development of asthma-like symptoms. By applying machine learning models, notably Random Forest (RF), I identified significant associations between exposure to chemicals such as hydrochloric acid and ammonia and the presence of respiratory symptoms in workers in the simulated dataset. Among the models tested, RF outperformed others, achieving the highest sensitivity and providing the most reliable predictions of symptomatic cases. RF has been widely used in occupational health studies to evaluate outcomes related to noise, dust, and chemical irritants *(26,29,30)*. The strong performance observed in this study supports its feasibility for predicting health outcomes related to chemical exposures. This aligns with previous research, reinforcing RF’s potential for use in occupational health risk modeling, particularly in laboratory environments with chemical irritants. In addition to identifying high-risk chemicals, the study found that workplace types (e.g., hospital vs. common areas) also played a critical role in determining asthma risk. These findings contribute to a deeper understanding of how routine cleaning practices in laboratories and medical facilities could inadvertently facilitate the development of asthma-like symptoms, particularly among vulnerable workers exposed to high levels of irritants.

Despite these key findings, the design of this simulated experiment presents several limitations that suggest avenues for future research. The heavily imbalanced data, reflecting the rare prevalence of occupational asthma *(28,31)*, constrained model techniques to fully disentangle more complex behavioral and environmental risk factors *(1,2,32)*. This highlights the need for further research using real-world data, which could better account for these interactions and improve model generalizability. Future studies could also explore more nuanced exposure data, including dose-response relationships or self-reported survey data *(7,33)*, to better capture gradations of risk and enhance model sensitivity (34)

This study provides a practical framework for evaluating chemical exposure risks in laboratory environments and emphasizes the importance of integrating machine learning models into occupational health and safety monitoring. This framework could be used to implement predictive models that identify high-risk workers in real-time, guiding safety interventions such as ventilation improvements or adjusted cleaning protocols. By identifying key chemical irritants, my research offers actionable insights that can directly inform lab safety protocols, particularly in areas where cleaning products are widely used. The findings underscore the need for regular monitoring of chemical exposures and suggest that predictive models could be incorporated into safety programs to better assess and mitigate health risks for laboratory workers. I believe extending this approach to other environmental health concerns, such as hazardous material handling, spills, or exposure to toxic substances, could further strengthen laboratory safety practices. Ultimately, integrating these insights into laboratory management could lead to improved occupational health standards and safer working conditions for researchers and staff.

## Data Availability

The synthetic dataset and code for this study are available on GitHub repository.
This repository includes:
01_asthma_data_simulation.py: Generates a synthetic cohort based on demographics and chemical exposures from Vizcaya et al. (2011).
02_classification_analysis.py: Implements machine learning models (Decision Tree, Random Forest, Gradient Boosting, XGBoost) with comprehensive metrics and evaluations.
The synthetic dataset and results for model analysis.
The data and code are fully reproducible and available for further research.

https://github.com/h-hedman/healthcare-data-science/tree/main/rf_asthma_lab

## Abbreviations

The following abbreviations are used in this manuscript:

DT: Decision tree
GB: Gradient Boosting
RF: Random Forest
XGB: XGBoost

